# Intentional injuries in Ethiopia: 2024–25 Ethiopia Demographic and Health Survey

**DOI:** 10.64898/2026.11.20.26368615

**Authors:** Awoke Mihretu, Mahder Dereb

## Abstract

**Background:** Injuries and violence disproportionately affect disadvantaged populations, particularly males, young people and those living in low- and middle-income countries, where about 90% of injury-related deaths occur. Preventing injuries is integral to achieving several Sustainable Development Goals. This study aimed to assess the magnitude, characteristics, spatial distribution, and individual- and household-level determinants of intentional injuries in Ethiopia.

**Methods:** We analysed the 2024–25 Ethiopia Demographic and Health Survey, a two-stage stratified cluster survey of 21,241 households (97,262 members) in 797 geo-referenced clusters. Household respondents reported injuries among members in the preceding 12 months; injuries due to violence/assault, self-harm or conflict were classified as intentional. Survey-weighted rates per 100,000 were estimated. Spatial patterns were assessed with global Moran’s I, Getis-Ord Gi^*^ hotspot analysis and Kulldorff’s Poisson scan statistic. Survey-weighted logistic regression compared intentional with unintentional non-traffic injuries (n = 222), and survey-weighted Poisson regression estimated household-level incidence rate ratios (IRRs).

**Results:** Of 435 injury events, 81 (18.6%) were intentional (38 conflict-related, 32 violence/assault, 11 self-harm), giving a weighted rate of 65.2 per 100,000 (95% CI 45.2–94.0) and a mortality rate of 14.6 per 100,000 (95% CI 7.5–28.3). Victims were mostly male (79.8%) and aged 15–29 years (51.0%), and gunshot was the commonest mechanism (36.7%). Intentional injuries were spatially clustered (Moran’s I = 0.053, p = 0.004); all 74 hotspots were in Tigray, Amhara and Afar, with the primary scan cluster in Afar (RR = 19.7, p = 0.001) and a secondary cluster in Tigray and northern Amhara (RR = 3.6, p = 0.009). Self-harm showed no clustering. Male sex (AOR = 3.33; 95% CI 1.25–8.89) and age 15–29 (AOR = 4.40; 1.27–15.21) and 30–44 years (AOR = 5.53; 1.35–22.66) were associated with an injury being intentional. Households in the northern conflict-affected regions had a six-fold higher rate (adjusted IRR = 6.44; 95% CI 2.91–14.25).

**Conclusions:** About one in five household-reported injuries in Ethiopia was intentional. These injuries mainly affected young men, often involved firearms, and were concentrated in conflict-affected regions. Self-harm was probably under-reported. Conflict resolution, firearm control, and trauma and psychosocial services targeted at these hotspots, together with better surveillance of suicide and self-harm, are needed.

## Background

Injuries are a leading cause of death and disability worldwide. Injuries and violence together cause about 4.4 million deaths each year, nearly 8% of all deaths, and violence-related injuries alone kill about 1.25 million people annually [1]. Among individuals aged 5–29 years, three of the five leading causes of death are injury-related: road traffic injuries, homicide, and suicide [1]. Intentional injuries are commonly grouped into self-directed violence (suicide and self-harm), interpersonal violence (assault and homicide) and collective violence (war and armed conflict) [2]. They fall disproportionately on men and young adults: worldwide, about four-fifths of homicide deaths and two-thirds of war deaths are among men [1].

Beyond physical harm and premature mortality, injuries and exposure to trauma can have lasting effects on mental health, substance use, chronic diseases, and social and economic well-being, perpetuating cycles of poverty and violence [1,3,4]. WHO’s recommended effective prevention requires addressing underlying individual, household, and societal risk factors through evidence-based, multisectoral interventions, alongside strengthening emergency trauma care, rehabilitation, and support services for survivors (1).

In LMICs, injuries constitute a major public health challenge, accounting for a substantial burden of premature mortality and disability, with road traffic injuries, interpersonal violence, and self-harm among the leading contributors [5]. In Ethiopia, the burden of injury remains a public health priority despite a long-term decline in injury mortality [6]. Global Burden of Disease estimates rank conflict and terrorism, interpersonal violence, and self-harm among the leading causes of injury-related death and long-term disability in the country, and they suggest that the burden from conflict and terrorism has not fallen as transport injuries and interpersonal violence have [6,7]. These estimates depend largely on modelling, because Ethiopia has no national injury surveillance system and only limited vital registration [6,7]. The available primary evidence comes mostly from hospital and small community studies, which rarely describe the population-level distribution of intentional injury.

Intentional injury is especially relevant in Ethiopia because of recent ongoing armed conflict and insecurity in Tigray, Amhara, Afar, and Oromia regions. The spread of illicit small arms and light weapons might be linked to rising homicide, robbery and kidnapping [8]. Suicide, the self-directed component of intentional injury, is probably under-recognized: the estimated national suicide rate is about 6 per 100,000 [9], but suicide is heavily stigmatized, and people who die by suicide may be denied normal funeral rites, which encourages concealment [10].

Preventing injuries is integral to achieving several Sustainable Development Goals (SDGs) (7). Accordingly, the Ethiopia Demographic and Health Survey (EDHS) first collected community-based injury data in 2016. Analyses of those data described the magnitude of road traffic injuries [11] and of unintentional injuries within households [12], but intentional injuries have not been examined separately, and their spatial distribution has not been studied using national data. The 2024–25 EDHS repeated the accident and injury module and, for the first time, recorded whether each non-traffic injury resulted from violence/assault, self-harm, or conflict [13]. Combined with geo-referenced cluster locations, this enables estimation of the population burden of intentional injuries and identification of where they are concentrated. This study aimed to assess the magnitude, characteristics, spatial distribution, and individual- and household-level determinants of intentional injuries in Ethiopia.

## Methods

### Study design and data collection Period

The study design was a community-based cross-sectional study. Data were collected between August 2024 and April 2025.

#### Data source

This study is a secondary analysis of the 2024–25 EDHS, the fifth nationally representative DHS in Ethiopia, implemented by the Ethiopian Statistical Service through the DHS Program [13].

#### Study population and sampling procedure

The survey used a two-stage stratified cluster sample: enumeration areas (clusters) were selected with probability proportional to size within 27 strata (each region divided into urban and rural areas), and households were then selected systematically within each cluster. The analysis used 21,241 interviewed households, with 97,262 household members, in 797 clusters with geographic coordinates.

### Data collection instruments and variables

The household questionnaire included an accident and injury module. The respondent was asked whether any household member had been injured or killed in an accident or injury in the 12 months before the survey. For each event, the survey recorded whether it was a road traffic accident; for non-traffic events, the mechanism (for example fall, gunshot, cut or stab, poisoning) and the circumstance (accidental, natural disaster, violence/assault, self-harm or conflict); the sex and age of the person; whether the person was still alive and, if not, whether the death was related to the injury; the types of injury sustained; and whether survivors continued to have health problems. The accident file (one record per event) was linked to the household file through the household identifier and to cluster coordinates through the cluster number.

Intentional injury was defined as a non-traffic injury whose circumstance was reported as violence/assault, self-harm or conflict. Unintentional non-traffic injuries were those reported as accidental, such as road traffic injuries, injuries from natural disasters and injuries of unknown circumstance.

Individual-level variables were the sex and age of the injured person (0–14, 15–29, 30–44, ≥45 years; current age for survivors and age at death for those who died). Household-level variables were place of residence (urban, rural), household wealth index (poor: poorest and poorer quintiles; middle; rich: richer and richest quintiles) and sex of the household head. Region was grouped as northern conflict-affected regions (Tigray, Afar, Amhara), central/southern regions (Oromia, Central Ethiopia, Sidama, South West Ethiopia, South Ethiopia) and other regions (Somali, Benishangul-Gumuz, Gambella, Harari, Addis Ababa, Dire Dawa). Regions were grouped because several had fewer than five intentional injuries; the northern grouping reflects the regions affected by armed conflict in the years before the survey [14].

#### Statistical analysis

All estimates accounted for the complex survey design using household sampling weights (hv005/1,000,000), clusters as primary sampling units (hv001) and sampling strata (hv023). Percentages are weighted and frequencies unweighted. Variances were estimated by Taylor linearization including all 797 clusters, so that clusters without an injury contributed to the variance. 95% confidence intervals (CIs) for proportions were logit-transformed. Injury and mortality rates per 100,000 household members were estimated as ratio estimators (weighted number of injured members divided by the weighted number of household members), with log-transformed 95% CIs.

In the spatial analysis, for each cluster, the numbers of injured and household members were computed. Global spatial autocorrelation of the cluster-level injury rate was assessed with Moran’s I, using row-standardised inverse-distance weights within 115 km (the smallest distance band that gave every cluster at least one neighbour), with significance tested by 999 random permutations. Local hot spots were identified with the Getis-Ord Gi^*^ statistic [15] using a fixed 115 km band; clusters with z > 1.96 (p < 0.05) and z > 2.58 (p < 0.01) were classified as hot spots. Purely spatial clusters with high rates were detected using Kulldorff’s discrete Poisson scan statistic [16] (cases = injured members; population = household members per cluster), with circular windows containing up to 50% of the population and 999 Monte Carlo replications. The most likely (primary) and non-overlapping secondary clusters with p < 0.05 are reported with relative risks (RR). The analyses were run for all intentional injuries and separately for conflict-related, violence/assault and self-harm injuries. The DHS Program randomly displaces cluster coordinates to protect confidentiality (up to 2 km in urban clusters and up to 5 km, with 1% up to 10 km, in rural clusters), and the displacement remains within the same zone [17].

Because intentional injuries were rare, we used two simple survey-weighted models. (1) Individual level: among injured persons with an intentional or unintentional non-traffic injury and known sex (n = 222; 79 intentional), survey-weighted logistic regression compared intentional with unintentional injuries, adjusting for sex, age, residence, wealth, region group and the sex of the household head. (2) Population level: among all 21,241 households, survey-weighted Poisson regression modelled the number of household members with an intentional injury, with the logarithm of household size as an offset, yielding incidence rate ratios (IRRs) for residence, wealth, region group and the sex of the household head. Both models used design-based (linearised, robust) standard errors, which are also valid under over-dispersion. Overall p-values for multi-category variables were obtained from design-adjusted Wald F tests. All statistical analyses were performed using Stata Statistical Software, Release 18. Anthropic Claude was used as an AI-assisted tool to support manuscript preparation and Python 3 code development for the analyses.

### Ethical considerations

This study used publicly available, de-identified EDHS data. The data were obtained with permission from The DHS Program after registration and approval of the analysis request (https://dhsprogram.com/data/) [18]. The EDHS protocol received ethical approval from the relevant national and ICF review bodies, and informed consent was obtained from respondents during data collection. No further ethical approval was required for this secondary analysis.

## Results

### Prevalence of intentional injuries

Of the 435 injury events reported among 97,262 household members, 81 (18.6%) were intentional: 38 conflict-related, 32 from violence or assault and 11 from self-harm. They occurred in 68 of the 797 clusters. The weighted rate of intentional injury was **65.2 per 100**,**000 household members** (95% CI 45.2–94.0) in the 12 months before the survey: 29.9 (18.1–49.1) for conflict-related injuries, 26.1 (13.7–49.6) for violence/assault and 9.2 (3.3–25.5) for self-harm. Twenty-four intentional injuries were fatal, an intentional-injury mortality rate of 14.6 per 100,000 (7.5–28.3), compared with 8.0 per 100,000 (3.2–19.8) for road traffic injuries, although the confidence intervals overlap (Table 1A).

**Table 1.**
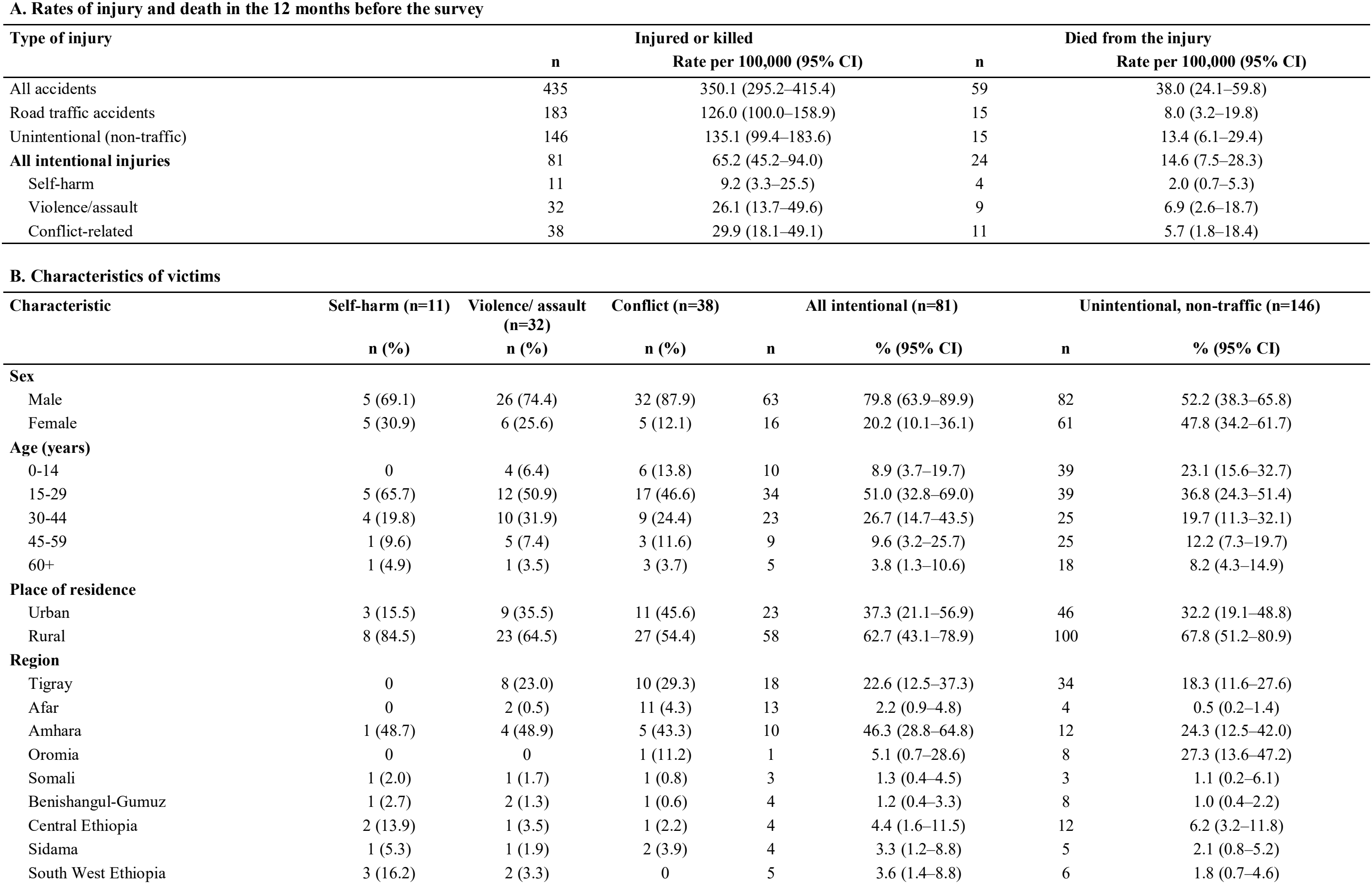
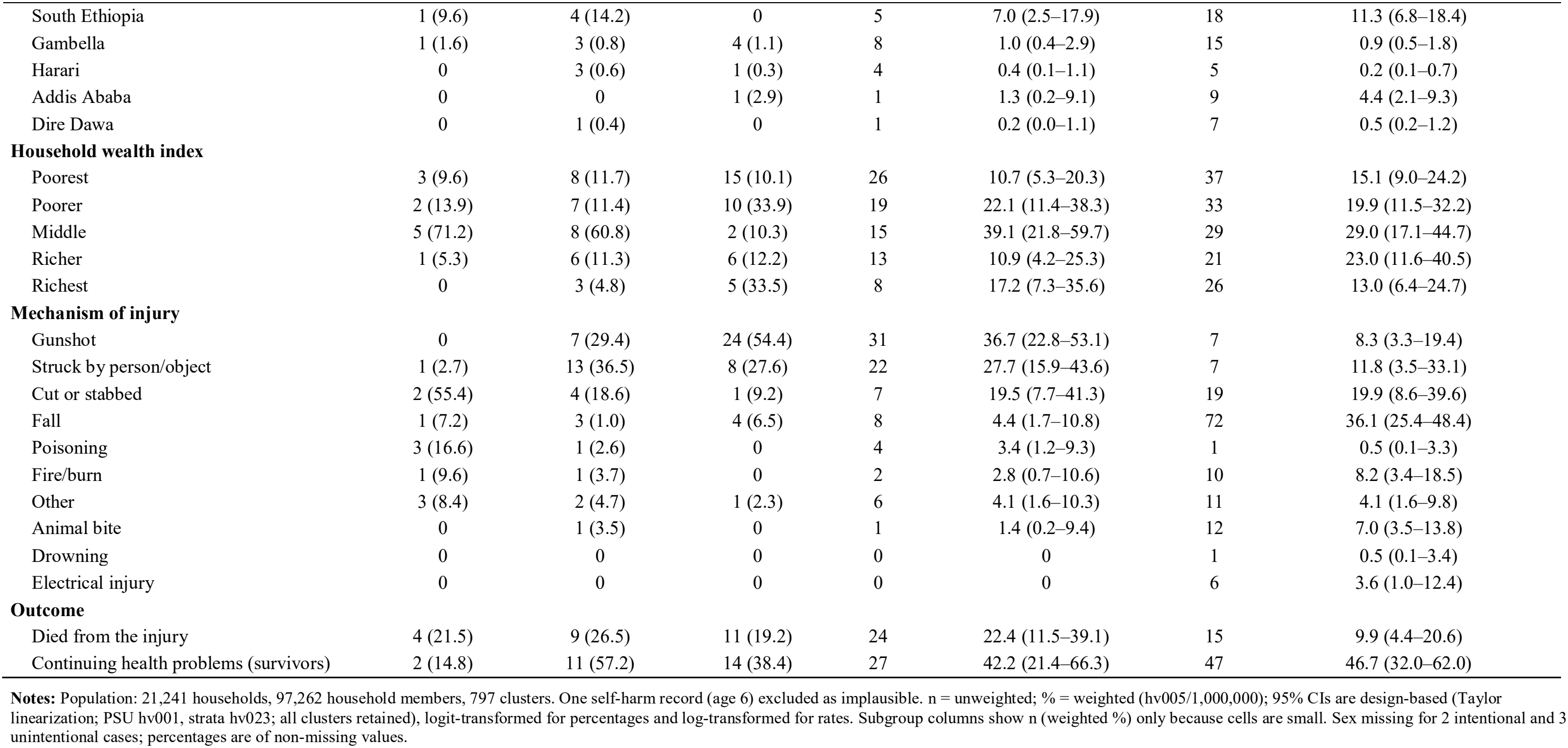
Intentional injuries in Ethiopia: (A) rates per 100,000 household members and (B) characteristics of victims, by intent, EDHS 2024–25.

| A. Rates of injury and death in the 12 months before the survey |  |  |  |  |  |  |  |
|---|---|---|---|---|---|---|---|
| Type of injury | Injured or killed |  | Died from the injury |  |  |  |  |
|  | n | Rate per 100,000 (95% CI) | n | Rate per 100,000 (95% CI) |  |  |  |
| All accidents | 435 | 350.1 (295.2–415.4) | 59 | 38.0 (24.1–59.8) |  |  |  |
| Road traffic accidents | 183 | 126.0 (100.0–158.9) | 15 | 8.0 (3.2–19.8) |  |  |  |
| Unintentional (non-traffic) | 146 | 135.1 (99.4–183.6) | 15 | 13.4 (6.1–29.4) |  |  |  |
| **All intentional injuries** | 81 | 65.2 (45.2–94.0) | 24 | 14.6 (7.5–28.3) |  |  |  |
| Self-harm | 11 | 9.2 (3.3–25.5) | 4 | 2.0 (0.7–5.3) |  |  |  |
| Violence/assault | 32 | 26.1 (13.7–49.6) | 9 | 6.9 (2.6–18.7) |  |  |  |
| Conflict-related | 38 | 29.9 (18.1–49.1) | 11 | 5.7 (1.8–18.4) |  |  |  |
| **B. Characteristics of victims** |  |  |  |  |  |  |  |
| **Characteristic** | **Self-harm (n=11)** | **Violence/ assault (n=32)** | **Conflict (n=38)** | **All intentional (n=81)** |  | **Unintentional, non-traffic (n=146)** |  |
|  | **n (%)** | **n (%)** | **n (%)** | **n** | **% (95% CI)** | **n** | **% (95% CI)** |
| **Sex** |  |  |  |  |  |  |  |
| Male | 5 (69.1) | 26 (74.4) | 32 (87.9) | 63 | 79.8 (63.9–89.9) | 82 | 52.2 (38.3–65.8) |
| Female | 5 (30.9) | 6 (25.6) | 5 (12.1) | 16 | 20.2 (10.1–36.1) | 61 | 47.8 (34.2–61.7) |
| **Age (years)** |  |  |  |  |  |  |  |
| 0-14 | 0 | 4 (6.4) | 6 (13.8) | 10 | 8.9 (3.7–19.7) | 39 | 23.1 (15.6–32.7) |
| 15-29 | 5 (65.7) | 12 (50.9) | 17 (46.6) | 34 | 51.0 (32.8–69.0) | 39 | 36.8 (24.3–51.4) |
| 30-44 | 4 (19.8) | 10 (31.9) | 9 (24.4) | 23 | 26.7 (14.7–43.5) | 25 | 19.7 (11.3–32.1) |
| 45-59 | 1 (9.6) | 5 (7.4) | 3 (11.6) | 9 | 9.6 (3.2–25.7) | 25 | 12.2 (7.3–19.7) |
| 60+ | 1 (4.9) | 1 (3.5) | 3 (3.7) | 5 | 3.8 (1.3–10.6) | 18 | 8.2 (4.3–14.9) |
| **Place of residence** |  |  |  |  |  |  |  |
| Urban | 3 (15.5) | 9 (35.5) | 11 (45.6) | 23 | 37.3 (21.1–56.9) | 46 | 32.2 (19.1–48.8) |
| Rural | 8 (84.5) | 23 (64.5) | 27 (54.4) | 58 | 62.7 (43.1–78.9) | 100 | 67.8 (51.2–80.9) |
| **Region** |  |  |  |  |  |  |  |
| Tigray | 0 | 8 (23.0) | 10 (29.3) | 18 | 22.6 (12.5–37.3) | 34 | 18.3 (11.6–27.6) |
| Afar | 0 | 2 (0.5) | 11 (4.3) | 13 | 2.2 (0.9–4.8) | 4 | 0.5 (0.2–1.4) |
| Amhara | 1 (48.7) | 4 (48.9) | 5 (43.3) | 10 | 46.3 (28.8–64.8) | 12 | 24.3 (12.5–42.0) |
| Oromia | 0 | 0 | 1 (11.2) | 1 | 5.1 (0.7–28.6) | 8 | 27.3 (13.6–47.2) |
| Somali | 1 (2.0) | 1 (1.7) | 1 (0.8) | 3 | 1.3 (0.4–4.5) | 3 | 1.1 (0.2–6.1) |
| Benishangul-Gumuz | 1 (2.7) | 2 (1.3) | 1 (0.6) | 4 | 1.2 (0.4–3.3) | 8 | 1.0 (0.4–2.2) |
| Central Ethiopia | 2 (13.9) | 1 (3.5) | 1 (2.2) | 4 | 4.4 (1.6–11.5) | 12 | 6.2 (3.2–11.8) |
| Sidama | 1 (5.3) | 1 (1.9) | 2 (3.9) | 4 | 3.3 (1.2–8.8) | 5 | 2.1 (0.8–5.2) |
| South West Ethiopia | 3 (16.2) | 2 (3.3) | 0 | 5 | 3.6 (1.4–8.8) | 6 | 1.8 (0.7–4.6) |
| South Ethiopia | 1 (9.6) | 4 (14.2) | 0 | 5 | 7.0 (2.5–17.9) | 18 | 11.3 (6.8–18.4) |
| Gambella | 1 (1.6) | 3 (0.8) | 4 (1.1) | 8 | 1.0 (0.4–2.9) | 15 | 0.9 (0.5–1.8) |
| Harari | 0 | 3 (0.6) | 1 (0.3) | 4 | 0.4 (0.1–1.1) | 5 | 0.2 (0.1–0.7) |
| Addis Ababa | 0 | 0 | 1 (2.9) | 1 | 1.3 (0.2–9.1) | 9 | 4.4 (2.1–9.3) |
| Dire Dawa | 0 | 1 (0.4) | 0 | 1 | 0.2 (0.0–1.1) | 7 | 0.5 (0.2–1.2) |
| **Household wealth index** |  |  |  |  |  |  |  |
| Poorest | 3 (9.6) | 8 (11.7) | 15 (10.1) | 26 | 10.7 (5.3–20.3) | 37 | 15.1 (9.0–24.2) |
| Poorer | 2 (13.9) | 7 (11.4) | 10 (33.9) | 19 | 22.1 (11.4–38.3) | 33 | 19.9 (11.5–32.2) |
| Middle | 5 (71.2) | 8 (60.8) | 2 (10.3) | 15 | 39.1 (21.8–59.7) | 29 | 29.0 (17.1–44.7) |
| Richer | 1 (5.3) | 6 (11.3) | 6 (12.2) | 13 | 10.9 (4.2–25.3) | 21 | 23.0 (11.6–40.5) |
| Richest | 0 | 3 (4.8) | 5 (33.5) | 8 | 17.2 (7.3–35.6) | 26 | 13.0 (6.4–24.7) |
| **Mechanism of injury** |  |  |  |  |  |  |  |
| Gunshot | 0 | 7 (29.4) | 24 (54.4) | 31 | 36.7 (22.8–53.1) | 7 | 8.3 (3.3–19.4) |
| Struck by person/object | 1 (2.7) | 13 (36.5) | 8 (27.6) | 22 | 27.7 (15.9–43.6) | 7 | 11.8 (3.5–33.1) |
| Cut or stabbed | 2 (55.4) | 4 (18.6) | 1 (9.2) | 7 | 19.5 (7.7–41.3) | 19 | 19.9 (8.6–39.6) |
| Fall | 1 (7.2) | 3 (1.0) | 4 (6.5) | 8 | 4.4 (1.7–10.8) | 72 | 36.1 (25.4–48.4) |
| Poisoning | 3 (16.6) | 1 (2.6) | 0 | 4 | 3.4 (1.2–9.3) | 1 | 0.5 (0.1–3.3) |
| Fire/burn | 1 (9.6) | 1 (3.7) | 0 | 2 | 2.8 (0.7–10.6) | 10 | 8.2 (3.4–18.5) |
| Other | 3 (8.4) | 2 (4.7) | 1 (2.3) | 6 | 4.1 (1.6–10.3) | 11 | 4.1 (1.6–9.8) |
| Animal bite | 0 | 1 (3.5) | 0 | 1 | 1.4 (0.2–9.4) | 12 | 7.0 (3.5–13.8) |
| Drowning | 0 | 0 | 0 | 0 | 0 | 1 | 0.5 (0.1–3.4) |
| Electrical injury | 0 | 0 | 0 | 0 | 0 | 6 | 3.6 (1.0–12.4) |
| **Outcome** |  |  |  |  |  |  |  |
| Died from the injury | 4 (21.5) | 9 (26.5) | 11 (19.2) | 24 | 22.4 (11.5–39.1) | 15 | 9.9 (4.4–20.6) |
| Continuing health problems (survivors) | 2 (14.8) | 11 (57.2) | 14 (38.4) | 27 | 42.2 (21.4–66.3) | 47 | 46.7 (32.0–62.0) |

**Notes:** Population: 21,241 households, 97,262 household members, 797 clusters. One self-harm record (age 6) excluded as implausible. n = unweighted; % = weighted (hv005/1,000,000); 95% CIs are design-based (Taylor linearization; PSU hv001, strata hv023; all clusters retained), logit-transformed for percentages and log-transformed for rates. Subgroup columns show n (weighted %) only because cells are small. Sex missing for 2 intentional and 3 unintentional cases; percentages are of non-missing values.

### Characteristics of victims

Intentional injury victims were predominantly male (79.8%; 95% CI 63.9–89.9), compared with 52.2% of unintentional injury victims, and half (51.0%) were aged 15–29 years (Table 1B). Most lived in rural areas (62.7%). In weighted terms, Amhara (46.3%) and Tigray (22.6%) accounted for over two-thirds of intentional injury victims. Afar contributed 13 of the 81 cases but a small weighted share, because the region is oversampled. Gunshot was the commonest mechanism (36.7%) and accounted for 54% of conflict-related injuries, followed by being struck by a person or object (27.7%) and cuts or stabbing (19.5%). Intentional injuries were more often fatal than unintentional non-traffic injuries (22.4% vs 9.9%), and 42.2% of survivors reported continuing health problems. Self-harm injuries (n = 11; 4 deaths) were spread across eight regions, and poisoning was the most frequently reported mechanism (3 of 11).

### Spatial distribution

Intentional injuries were spatially clustered across Ethiopia (Global Moran’s I = 0.053, z = 4.35, p = 0.004) (Table 2). Getis-Ord Gi^*^ identified 74 hot-spot clusters at the 95% level, all in Tigray (31), Amhara (27) and Afar (16) (Figure 1B). The scan statistic detected a primary cluster of six DHS clusters in Afar centred at 10.39°N, 40.71°E (radius 56 km), with 10 observed cases against 0.6 expected (RR = 19.7, p = 0.001). A secondary cluster covered 87 DHS clusters in Tigray and northern Amhara (24 observed vs 8.6 expected; RR = 3.6, p = 0.009) (Figure 1C).

**Table 2.** Spatial autocorrelation and spatial scan clusters of intentional injuries, by intent, EDHS 2024–25.

| Outcome | Cases / clusters with case | Global Moran's I (z); p | Scan cluster | Centre; radius | Regions (number of DHS clusters) | DHS clusters | Observed / expected | RR | LLR; p |
|---|---|---|---|---|---|---|---|---|---|
| All intentional injuries | 81 / 68 | 0.053 (4.35); 0.004 | Primary | 10.39°N, 40.71°E; 56 km | Afar (6) | 6 | 10 / 0.6 | 19.7 | 19.7; 0.001 |
|  |  |  | Secondary | 14.11°N, 36.95°E; 303 km | Tigray (61), Amhara (25), Afar (1) | 87 | 24 / 8.6 | 3.6 | 11.0; 0.009 |
| Conflict-related | 38 / 32 | 0.067 (5.31); 0.005 | Primary | 10.39°N, 40.71°E; 56 km | Afar (6) | 6 | 9 / 0.3 | 43.5 | 24.0; 0.001 |
| Violence/assault | 32 / 29 | 0.017 (1.42); 0.095 | Primary | 14.41°N, 37.77°E; 218 km | Tigray (55), Amhara (10), Afar (1) | 66 | 11 / 2.5 | 6.1 | 9.1; 0.038 |
| Self-harm | 11 / 11 | -0.001 (0.02); 0.978 | None significant | — | — | — | — | — | most likely: p=0.149 |

**Notes:** Global Moran's I on cluster-level injury rate per 1,000 household members, inverse-distance weights within 115 km, 999 permutations. Spatial scan: Kulldorff discrete Poisson model, circular windows up to 50% of the population, 999 Monte Carlo replications (smallest attainable p = 0.001). RR = relative risk inside vs outside the window; LLR = log-likelihood ratio.

**Figure 1.**
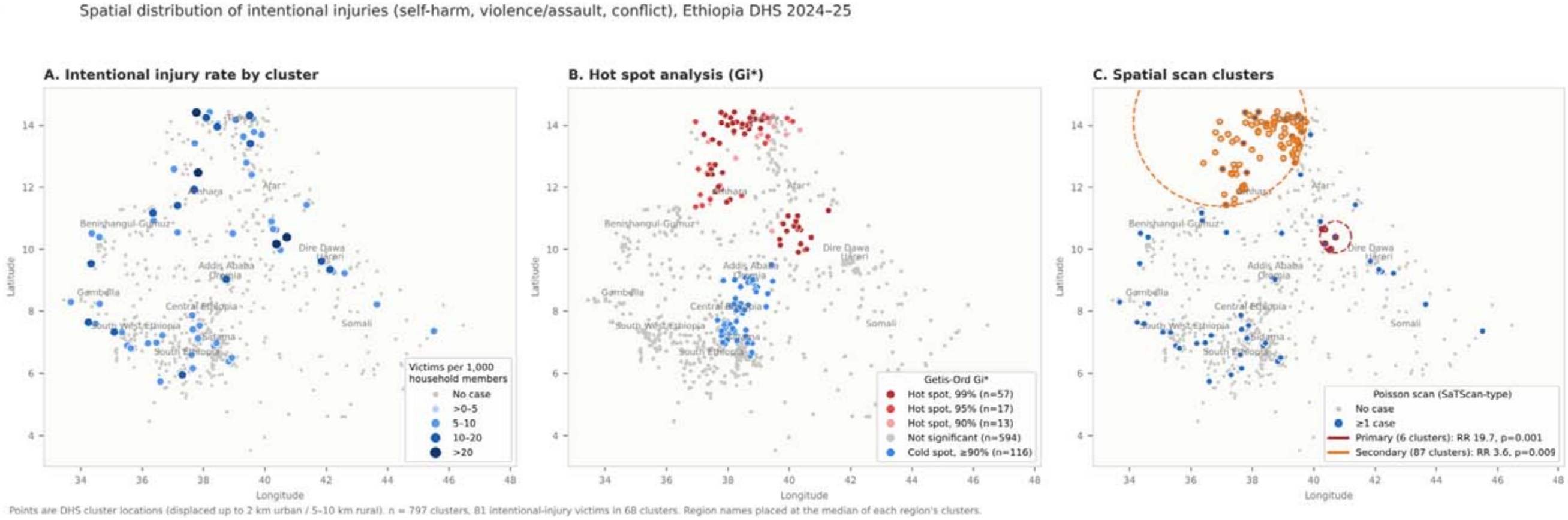
Spatial distribution of intentional injuries: (A) cluster injury rate, (B) Getis-Ord Gi^*^ hot spots, (C) spatial scan clusters, EDHS 2024–25.

By intent, conflict-related injuries showed the strongest clustering (Moran’s I = 0.067, p = 0.005), with a primary scan cluster in the same area of Afar (9 observed vs 0.3 expected; RR = 43.5, p = 0.001). Violence/assault injuries did not show significant global autocorrelation (Moran’s I = 0.017, p = 0.095) but formed a significant scan cluster in Tigray and northern Amhara (66 DHS clusters; RR = 6.1, p = 0.038). Self-harm injuries showed no spatial clustering (Moran’s I = −0.001, p = 0.978; most likely scan cluster p = 0.149) (Figure 2).

**Figure 2.**
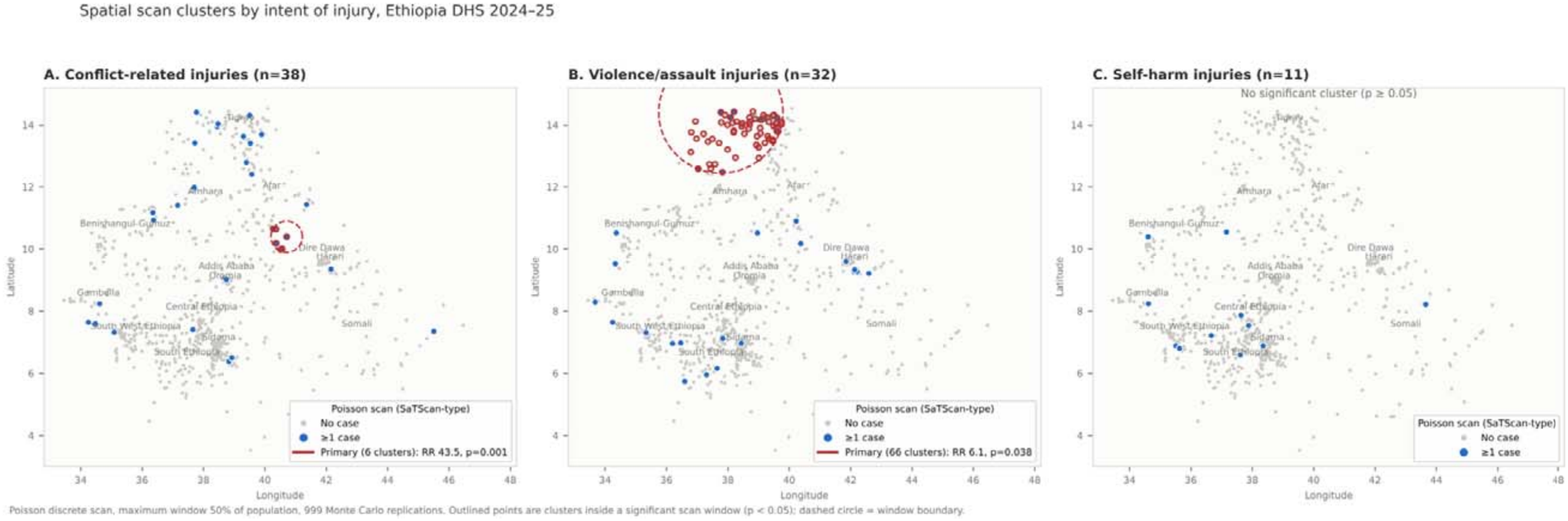
Spatial scan clusters for conflict-related, violence/assault and self-harm injuries, EDHS 2024–25.

### Factors associated with intentional injury

Among 222 injured persons (79 intentional injuries in 172 clusters), males had higher odds of an injury being intentional than females (AOR = 3.33; 95% CI 1.25–8.89). Compared with children aged 0–14 years, the odds were higher among those aged 15–29 (AOR = 4.40; 1.27–15.21) and 30–44 years (AOR = 5.53; 1.35–22.66). Injured persons in the northern conflict-affected regions had about three times the odds of their injury being intentional compared with those in central/southern regions (AOR = 2.96; 1.04–8.39), although the overall test for region group was not significant (p = 0.099). Place of residence, household wealth and sex of the household head were not associated with an injury being intentional **(Table 3)**.

**Table 3.** Individual-level factors associated with an injury being intentional rather than unintentional (non-traffic) among injured household members, EDHS 2024–25.

| Variable | Intentional / total (n) | Weighted % intentional | Crude OR (95% CI) | Adjusted OR (95% CI) | p |
|---|---|---|---|---|---|
| **Sex of injured person** |  |  |  | overall p = 0.016 |  |
| Female | 16/77 | 17.1 | 1.00 | 1.00 |  |
| Male | 63/145 | 42.8 | 3.63 (1.42–9.29) | 3.33 (1.25–8.89) | 0.016 |
| **Age (years)** |  |  |  | overall p = 0.045 |  |
| 0-14 | 10/48 | 15.9 | 1.00 | 1.00 |  |
| 15-29 | 32/71 | 39.9 | 3.51 (1.02–12.05) | 4.40 (1.27–15.21) | 0.019 |
| 30-44 | 23/47 | 40.2 | 3.55 (0.95–13.19) | 5.53 (1.35–22.66) | 0.018 |
| 45+ | 14/56 | 24.6 | 1.72 (0.46–6.45) | 2.24 (0.53–9.49) | 0.272 |
| **Place of residence** |  |  |  | overall p = 0.279 |  |
| Urban | 22/66 | 36.5 | 1.00 | 1.00 |  |
| Rural | 57/156 | 31.0 | 0.78 (0.27–2.27) | 0.38 (0.07–2.18) | 0.279 |
| **Household wealth** |  |  |  | overall p = 0.653 |  |
| Poor (poorest, poorer) | 44/113 | 31.4 | 1.00 | 1.00 |  |
| Middle | 14/43 | 39.3 | 1.41 (0.49–4.09) | 0.80 (0.28–2.28) | 0.680 |
| Rich (richer, richest) | 21/66 | 27.9 | 0.85 (0.30–2.40) | 0.44 (0.08–2.57) | 0.362 |
| **Region group** |  |  |  | overall p = 0.099 |  |
| Central/southern (Oromia, Central, Sidama, South West, South Ethiopia) | 19/66 | 19.2 | 1.00 | 1.00 |  |
| North (Tigray, Afar, Amhara) | 40/89 | 44.6 | 3.39 (1.27–9.08) | 2.96 (1.04–8.39) | 0.042 |
| Other (Somali, Benishangul-Gumuz, Gambella, Harari, Addis Ababa, Dire Dawa) | 20/67 | 23.5 | 1.29 (0.42–3.99) | 1.34 (0.38–4.73) | 0.650 |
| **Sex of household head** |  |  |  | overall p = 0.979 |  |
| Male | 49/146 | 33.7 | 1.00 | 1.00 |  |
| Female | 30/76 | 31.0 | 0.89 (0.31–2.50) | 0.98 (0.31–3.09) | 0.979 |

**Notes:** n = 222 injured persons (79 intentional, 143 unintentional) in 172 clusters; 5 persons with missing sex excluded. Survey-weighted logistic regression with design-based (linearized) standard errors; adjusted model includes all variables shown. p = Wald test for each category; overall p = design-adjusted Wald F test. About 8 events per variable; estimates are imprecise.

Among all 21,241 households, the weighted intentional-injury rate was 154.9 per 100,000 household members in the northern regions, compared with 25.8 in central/southern regions. After adjustment, the rate in the northern regions was more than six times higher (IRR = 6.44; 95% CI 2.91–14.25; p < 0.001); the other regions did not differ from central/southern regions (IRR = 1.01; 0.37–2.73). In the adjusted model, rural residence (IRR = 0.19; 0.05–0.72) and rich households (IRR = 0.25; 0.07–0.82) had lower rates. These associations were not present in the crude analysis (rural IRR = 0.67; rich IRR = 0.85), which suggests that they partly reflect the strong correlation between wealth and urban residence, so they should be interpreted with caution. Sex of the household head was not associated with the rate (IRR = 1.31; 0.64–2.69) **(Table 4)**.

**Table 4.** Household-level rates and incidence rate ratios (IRR) of intentional injury among all households, EDHS 2024–25.

| **Household characteristic** | **Intentional injuries (n)** | **Household members (n)** | **Weighted rate per 100,000** | **Crude IRR (95% CI)** | **Adjusted IRR (95% CI)** | **p** |
|---|---|---|---|---|---|---|
| **Place of residence** |  |  |  |  | overall p = 0.015 |  |
| Urban | 23 | 29,374 | 85.7 | 1.00 | 1.00 |  |
| Rural | 58 | 67,888 | 57.1 | 0.67 (0.30–1.50) | 0.19 (0.05–0.72) | 0.015 |
| **Household wealth** |  |  |  |  | overall p = 0.020 |  |
| Poor (poorest, poorer) | 45 | 48,197 | 53.7 | 1.00 | 1.00 |  |
| Middle | 15 | 15,619 | 127.5 | 2.38 (0.99–5.70) | 2.13 (0.94–4.86) | 0.071 |
| Rich (richer, richest) | 21 | 33,446 | 45.6 | 0.85 (0.38–1.90) | 0.25 (0.07–0.82) | 0.023 |
| **Region group** |  |  |  |  | overall p = <0.001 |  |
| Central/southern (Oromia, Central, Sidama, South West, South Ethiopia) | 19 | 40,321 | 25.8 | 1.00 | 1.00 |  |
| North (Tigray, Afar, Amhara) | 41 | 22,266 | 154.9 | 5.99 (2.81–12.78) | 6.44 (2.91–14.25) | <0.001 |
| Other (Somali, Benishangul-Gumuz, Gambella, Harari, Addis Ababa, Dire Dawa) | 21 | 34,675 | 32.7 | 1.27 (0.53–3.03) | 1.01 (0.37–2.73) | 0.988 |
| **Sex of household head** |  |  |  |  | overall p = 0.459 |  |
| Male | 51 | 72,017 | 58.4 | 1.00 | 1.00 |  |
| Female | 30 | 25,245 | 90.0 | 1.54 (0.78–3.05) | 1.31 (0.64–2.69) | 0.459 |

**Notes:** Survey-weighted Poisson regression of the number of household members with an intentional injury (81 injuries in 79 of 21,241 households), offset = log(household size), design-based (robust) standard errors; adjusted model includes all variables shown. Rates are weighted per 100,000 household members.

## Discussion

This study provides the first national, population-based estimates of intentional injuries in Ethiopia that separate conflict-related, interpersonal and self-directed violence, together with their spatial distribution. About one in five injuries reported by households in the year before the survey was intentional, giving a rate of 65 per 100,000 household members and 15 deaths per 100,000. Intentional injuries were concentrated among young adult men, were dominated by gunshot wounds, were more than twice as often fatal as unintentional non-traffic injuries, and were strongly clustered in Afar, Tigray and northern Amhara. Self-harm was the smallest component and showed no geographic pattern.

Another important finding is the geographic concentration of intentional injuries in the north and north-east. The primary scan cluster, in Afar around 10.4°N, 40.7°E, coincides with zone 3 of Afar (Amibara and Gewane woredas), the centre of the long-running Afar–Somali (Issa) border dispute, where contested kebeles have repeatedly been the scene of deadly armed clashes [14]. Within this small area, conflict-related injuries were more than forty times more frequent than expected. The secondary cluster in Tigray and northern Amhara corresponds to the areas affected by the 2020–2022 northern Ethiopia war, by subsequent clashes in disputed territories of southern Tigray, and by the ongoing Fano insurgency in Amhara, which has been marked by violence against civilians. That gunshot accounted for more than half of conflict-related injuries is consistent with the reported proliferation of illicit small arms and light weapons in Ethiopia [8]. These findings support the Global Burden of Disease observation that conflict and terrorism remain a major, persistent cause of injury in Ethiopia [6], and they show that household surveys can capture this burden at the population level, which facility-based data from conflict areas, where services are disrupted, often cannot.

Violence/assault injuries, mainly being struck by a person or object and gunshots, also clustered in Tigray and northern Amhara, although less strongly. In post-conflict and insecure settings, the line between collective and interpersonal violence is blurred: widespread weapons, displacement, weakened policing and economic disruption are all recognized drivers of homicide, robbery and kidnapping [8]. The overlap of the violence and conflict clusters suggests that the respondents’ classification of an event as ‘conflict’ or ‘violence’ may sometimes reflect the same underlying insecurity, and that prevention in these areas needs to address both.

The self-harm death rate in this study (2.0 per 100,000) is about one-third of the modelled national suicide mortality rate of about 6 per 100,000 [9], and the overall self-harm rate (9 per 100,000, fatal and non-fatal combined) is far below the 12-month prevalence of suicide attempts of about 4% reported in a community study in rural Ethiopia [10]. Under-reporting is the most likely explanation. Suicide and self-harm are stigmatised in Ethiopia and are often considered sinful [10]; in a household interview, a family member may be unwilling to describe an injury as self-inflicted and may report it as accidental instead. Our self-harm estimate should therefore be regarded as a minimum, and the household injury module is not a suitable tool for measuring suicide. The absence of spatial clustering for self-harm should also be interpreted cautiously, because 11 cases provide little statistical power.

On the associated factors, male sex and young adulthood (15–44 years) were the strongest individual-level factors distinguishing intentional from unintentional injuries, consistent with the global pattern in which most victims of homicide and war are young men [1]. Young men are more likely to be combatants, to carry weapons and to be exposed to violence outside the home. At the population level, living in the northern conflict-affected regions was associated with a six-fold higher rate of intentional injury, independent of residence, wealth and sex of the household head. Household wealth and residence showed no consistent association: neither was associated in crude analyses, and the adjusted estimates were imprecise. This suggests that in the current Ethiopian context, intentional injury is driven more by place, and the insecurity that comes with it, than by household socioeconomic position.

### Strengths and limitations

The main strengths of this study are its use of the most recent nationally representative data, the separation of intentional injuries by intent, a design-based analysis that includes all clusters, and the combination of global, local and scan-based spatial methods, which gave consistent results. Several limitations should be considered. First, injuries were reported by a single household respondent for all members over a 12-month recall period. Household injury surveys in sub-Saharan Africa show that rates fall substantially with longer recall, especially for minor injuries, whereas severe injuries are recalled more reliably [19,20]. Second, the intent of each injury was reported by the respondent, not verified, and stigma probably led to under-reporting of self-harm and possibly misclassification between conflict and interpersonal violence. Third, households living in areas that were inaccessible because of insecurity, and people who were displaced, may be under-represented, which would bias the estimates for conflict-affected areas downwards. Fourth, the number of intentional injuries was small (81), so several estimates, particularly by region and in the regression models, are imprecise, and the regression results should be regarded as exploratory. We used simple survey-weighted models because multilevel models were unstable with so few events. Fifth, cluster coordinates are displaced by up to 2–10 km, which can blur small-area patterns, although it is unlikely to change clusters of the size detected here [17].

## Conclusion

About one in five household-reported injuries in Ethiopia in 2024–25 was intentional, at a rate of 65 per 100,000 population. Most victims were young men, gunshot wounds were common, and these injuries were strongly concentrated in the conflict-affected areas of Afar, Tigray and northern Amhara. Self-harm was rare in the data, most likely because it is under-reported. Psychosocial care, firearm control and trauma care, together with better surveillance of suicide and self-harm, are needed to reduce the burden of intentional injury in Ethiopia.

## Data Availability

DHS data

## Declarations

Ethics approval and consent to participate: Not applicable for this secondary analysis. The study used publicly available, de-identified data from the DHS Program, with no interaction between the researchers and the participants; permission to use the data was obtained from the DHS Program. The original 2024–25 EDHS data collection was approved by the Ethiopian Public Health Institute Institutional Review Board and the ICF Institutional Review Board, and informed consent was obtained from all participants by the survey implementers.

## Responsible AI use

Claude (Anthropic) assisted with data management and analysis code, and with drafting the manuscript. The authors reviewed and verified all analyses, results and text.

## Consent for publication

Not applicable.

Availability of data and materials: The datasets are available from the DHS Program (https://dhsprogram.com) on registration. Analysis code, including the record-linkage procedure, is provided in Additional file 2.

## Authors’ contributions

AM conceptualised and drafted the manuscript. MD conceptualised the study and led the data analysis. Both authors read and approved the final manuscript.

## Acknowledgements

We thank the DHS Program and the Ethiopian Statistical Service for access to the data, and the families who took part in the survey.

